# A novel framework for quantifying the clinical impact of substandard and falsified antimicrobials: application to typhoid fever

**DOI:** 10.1101/2025.07.08.25331006

**Authors:** Sean Cavany, Lawrence Adipo, Kerlijn van Assche, Celine Caillet, Christiane Dolecek, Cathrin Hauk, Jessica Mendes, Stella Nanyonga, Nicole Stoesser, Paul N. Newton, Christopher M. Parry, Ben S. Cooper

**Affiliations:** Drug Resistant Infections and Disease Dynamics (DRIaDD) research group, NDM Centre for Global Health Research, Nuffield Department of Medicine, University of Oxford, Oxford, OX3 7LF; Medicine Quality Research Group (MQRG), NDM Centre for Global Health Research, Nuffield Department of Medicine, University of Oxford, Oxford, OX3 7LF; Infectious Diseases Data Observatory, NDM Centre for Global Health Research, Nuffield Department of Medicine, University of Oxford, Oxford, OX3 7LF; Mahidol Oxford Research Unit, Faculty of Tropical Medicine, Mahidol University, Bangkok; Antimicrobial resistance, antimicrobial consumption and burden estimation (MICRoBE) research group, NDM Centre for Global Health Research, Nuffield Department of Medicine, University of Oxford, Oxford, OX3 7LF; Modernizing Medical Microbiology, Nuffield Department of Medicine, University of Oxford, Oxford, OX3 9DU; NIHR Health Protection Research Unit in Healthcare Associated Infections and Antimicrobial Resistance at University of Oxford in partnership with Public Health England, Nuffield Department of Medicine, Oxford, Oxford, OX3 9DU; NIHR Oxford Biomedical Research Centre, Oxford University Hospitals NHS Foundation Trust, John Radcliffe Hospital, Oxford, Oxford, OX3 7LF; Centre for Tropical Medicine and Global Health, Nuffield Department of Medicine, University of Oxford

## Abstract

**Objectives:** Substandard and falsified (SF) medicines are a neglected public health threat, with an estimated 7% of antimicrobials in low- and middle-income countries SF. However, quantifying their clinical impact remains challenging. We developed a general framework for estimating population-level impacts of SF antimicrobials on patient outcomes and apply it to SF fluoroquinolones in typhoid treatment.

**Patients and methods:** The framework combines two data sources: published surveys of antimicrobial quality to characterize the distribution of active pharmaceutical ingredient (API) content and individual-level patient data to estimate dose-response relationships. These are synthesized to evaluate population-level impacts. We extracted data from surveys of fluoroquinolone quality and fitted generalized additive models to data from seven clinical trials to estimate the causal effect of ofloxacin dose on typhoid outcomes.

**Results:** Application to typhoid demonstrated that while dose had negligible overall effect within trial dose ranges, lower doses worsened outcomes for non-susceptible strains. When the minimum inhibitory concentration was 1 mg/L, ofloxacin matching observed ofloxacin quality (mean %API=102.8%) improved fever clearance by 4.1 hours (90% CrI: 0.75–8.7) compared to 100% API. At the same minimum inhibitory concentration, ofloxacin whose %API instead matched the ciprofloxacin surveys (mean %API=93.6%) worsened fever clearance by 11 hours (90% CrI: 1.9– 25).

**Conclusions:** This framework can quantify the impact of SF antimicrobials on different pathogen– antimicrobial pairs. For typhoid, it demonstrates that SF fluoroquinolones likely have a substantial impact on treatment outcomes in settings like South Asia where non-susceptible strains are prevalent. Increased vigilance around antimicrobial quality is important in such settings.

## Introduction

Enteric fever (typhoid and paratyphoid fever, caused by *Salmonella enterica* serotypes Typhi and Paratyphi A respectively) caused an estimated 9,300,000 cases and 110,000 deaths in 2021.^1^ This epidemiological burden is concentrated in sub-Saharan Africa and South & Southeast Asia, and among children and young adults. Efficacious treatments exist for drug-susceptible typhoid. The WHO AWaRe book^2^ recommends ciprofloxacin for 7–10 days when the risk of fluoroquinolone resistance is low. Following a prompt course of treatment, most patients will recover, but without efficacious treatment, typhoid can cause a range of severe symptoms, including gastrointestinal haemorrhage and intestinal perforation, or death.^3^ Despite the existence of such efficacious treatments, they may be undermined by several factors. One such factor is rising levels of fluoroquinolone resistance^4^ which is associated with worse treatment outcomes.^5^ Another, currently under-explored, factor potentially undermining treatment is the use of substandard and falsified (SF) antibiotics.

This challenge exemplifies a broader problem affecting many infectious diseases: while SF antibiotics constitute approximately 7% of all antibiotics in low- and middle-income countries (LMICs),^6^ their clinical impact remains poorly quantified. Substandard medical products are “authorized medical products that fail to meet either their quality standards or their specifications, or both” and falsified medical products are those which “deliberately/fraudulently misrepresent their identity, composition or source”.^7,8^ SF medical products can contain the wrong concentration of active pharmaceutical ingredient (API) and occasionally they may contain no API at all. They may also have poor bioavailability. However, how different API concentrations affect treatment outcomes remains unclear for most antimicrobials. Previous studies have attempted to quantify the public health impact of SF antimicrobials on childhood pneumonia^6^ and malaria,^6,9^ finding a potentially large association between SF medicines and mortality. However, these studies did not use individual patient data and had to assume rather than estimate how much SF antimicrobials worsen the case fatality risk or drug efficacy. To date, the role of SF antimicrobials in typhoid outcomes has not been quantified. More broadly, no general framework exists for quantifying the clinical impact of SF antimicrobials across different pathogen-antimicrobial combinations.

To address this broader knowledge gap, we aimed to develop a general framework to quantify the population-level impacts of SF antimicrobials on clinical outcomes. We then applied this framework to quantify the effect of SF fluoroquinolones on typhoid outcomes. The framework has three steps (Fig 1). First, we characterize the distribution of %API (relative to that stated on the packaging) reported in published random surveys of the relevant antimicrobial. Second, we estimate the causal relationship between antimicrobial dose and clinical outcomes by reanalyzing individual-level data from clinical trials. Third, we synthesize the results of the first two steps to quantify the potential population-level effect of SF antimicrobials. This approach can be used for any antimicrobial-pathogen combinations and can be updated as new data become available.

**Fig 1.**
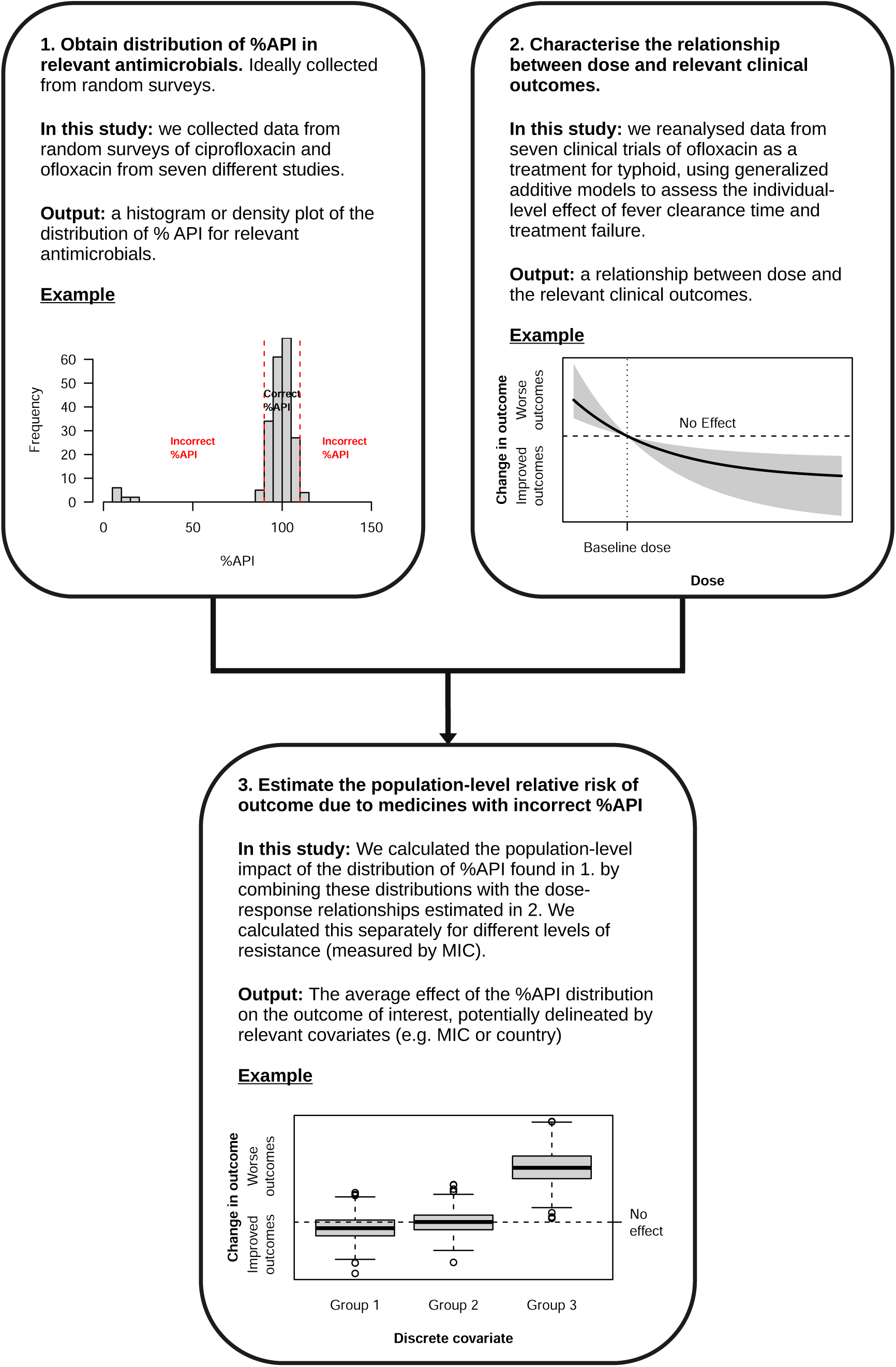
Overview of the framework.

## Patients and methods

### Data

#### Substandard and falsified antimicrobial data

We focus on two fluoroquinolone antibiotics: ciprofloxacin, which is the first line treatment for typhoid fever recommended by the WHO AWaRe book,^2^ and ofloxacin, which is the treatment used in the included clinical trials. We extracted data from the medicine quality surveyor (https://www.iddo.org/mqsurveyor/).^10^ Included studies used random sampling of the health facilities and reported the results of a %API assay for ciprofloxacin and/or ofloxacin.^11^

#### Typhoid clinical trial data

We use data from seven randomized clinical trials conducted across three sites in Vietnam between 1992 and 2001;^12–18^ see Table S1 for additional details about each of the included studies. Each of these trials either compared different durations of ofloxacin treatment or compared ofloxacin treatment to treatment with another antibiotic or combination of antibiotics (cefixime, ceftriaxone, azithromycin, or a combination of ofloxacin & azithromycin). We only use data from the ofloxacin arms of these trials. Note that six of these seven trials typically explored a shorter duration of treatment than the seven days recommended by the AWaRe book—see Table S1 for more details.

Fever clearance time was defined as the time in hours from when an individual started treatment to when their temperature decreased and remained at or below 37.5°C for 48h. Treatment failure was defined as any of four possible outcomes: a. the persistence of fever and at least one other symptom of typhoid for more than 7 days following the start of treatment, or b. the development of severe complications during or after enrolment, or c. the isolation of *Salmonella enterica* serovar Typhi from blood after the completion of treatment, or d. relapse, defined as recurrence of symptoms and signs suggestive of typhoid fever within four weeks following discharge.

In two studies,^16,18^ ofloxacin was administered as a fixed dose of 400mg/day, in two divided doses. In these two cases, it was possible to work out precisely the patient bodyweight-adjusted dose in mg/kg/day, 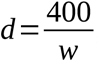, where *w* is each individual’s weight in kg. In the remaining five studies,^12–15,17^ ofloxacin was administered according to a target dose of *t* = 10, 15, or 20 mg/kg/day. In these cases, we estimated the actual weight-adjusted dose received by assuming the standard formulation of 200mg tablets was used^19^ and those tablets were administered in half-tablet increments, consistent with standard trial procedures. Hence, we estimated the actual weight-adjusted dose as:

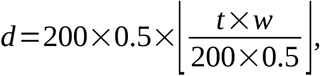

where ⌊ *x* ⌋ is a function that rounds *x* down to the nearest integer.

### Modelling

We focussed on two clinical outcomes of interest: time to fever clearance (h) and treatment failure probability To investigate the causal effect of different doses on the outcomes, we began by developing a directed acyclic graphs (DAG) which represents assumed causal relationships among variables based on subject matter expertise and existing literature (Fig. 2). We used the *dagitty*^20^ package (version 0.3-4) in *R*^21^ to analyse this DAG. We tested the DAG’s implied conditional independencies using linear models; all were confirmed except for a weak dose–MIC association in one trial. This DAG implies that to understand the causal effect of dose on outcome, we need an adjusted model that includes covariates representing study ID and patient bodyweight. We adjusted for study ID by using a model with a random intercept for study ID. We also included MIC in the model, as we were interested in the interaction between MIC and dose in affecting the outcomes. We excluded 34/572 (5.9%) of patients for whom MIC was not recorded.

**Fig 2:**
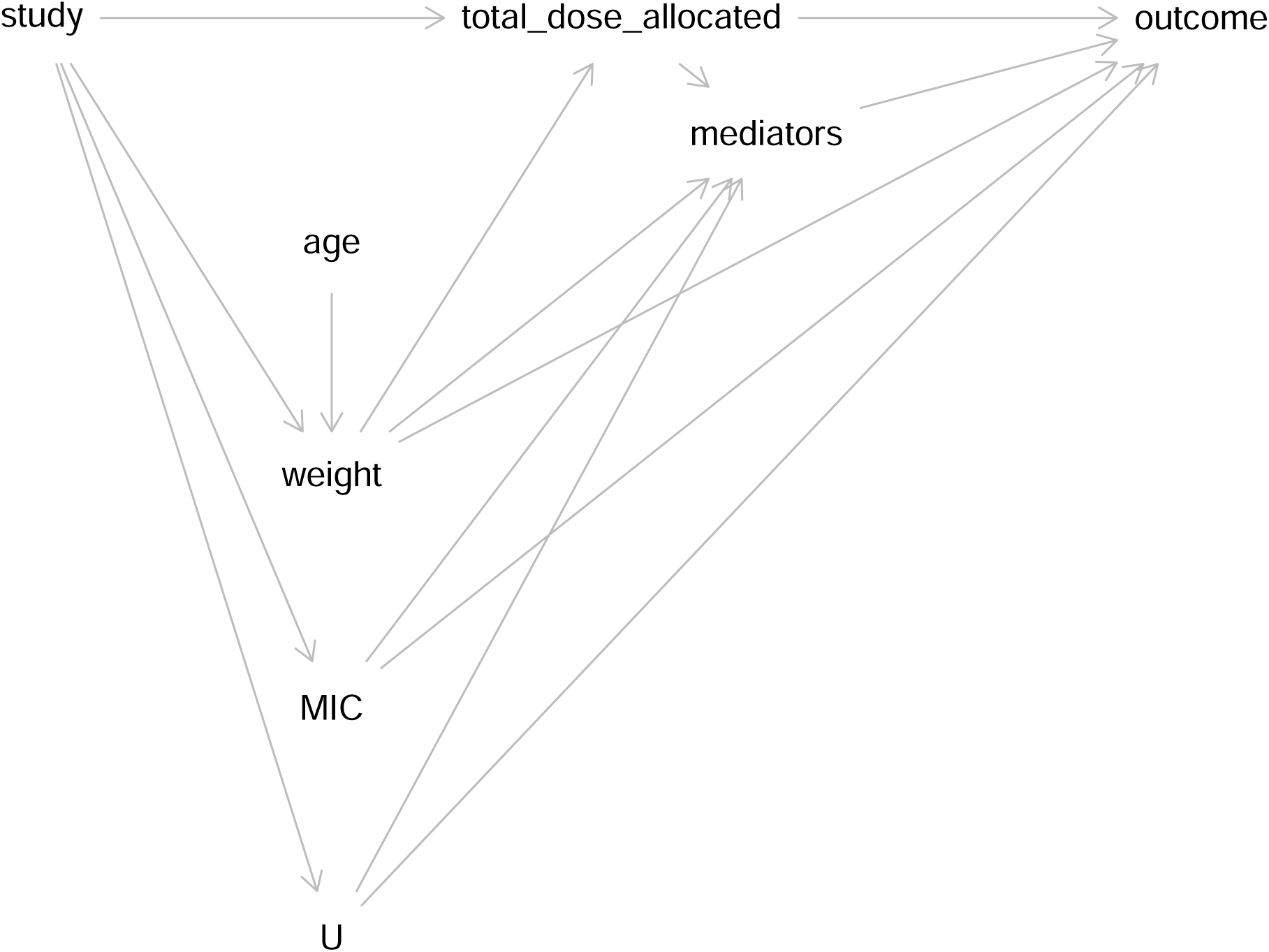
Directed acyclic graph showing the assumed causal relationships between key variables in our data. U represents unmeasured potential confounders, and mediators represents mediating variables which may or may not be measured. Outcome is either fever clearance time or treatment failure. MIC, minimum inhibitory concentration

To simplify the analysis, we dealt with the total bodyweight-adjusted dose (mg/kg) received by patients, which is the product of the daily dose (mg/kg/day) and duration of treatment (days). We used generalized additive models to allow for smooth data-driven relationships between the variables and the outcome, though this framework could naturally accommodate many other modelling approaches within a causal inference framework. We fitted these models to the trial data in a Bayesian workflow using Markov chain Monte Carlo, using the *brms* package^22–24^ (version 1.9-1) in *R*^21^. Given the considerations above, our baseline model was then:

`outcome ∼ t2(total_dose_allocated, MIC) + s(weight) + s(study, bs = “re”)` where `t2()` represents a tensor (interaction) term, s() represents a spline term, and `bs = “re“` indicates a random effect of study. Here outcome is either time to fever clearance (h) or the failure probability. For time to fever clearance we used a gamma likelihood and an inverse link, and for failure probability we used a binomial likelihood and logit link. We used weakly informative priors on all parameters: in fever clearance time models, we used Normal(5, 1) for the intercept, Normal(0, 0.5) for the spline standard deviations, and Gamma(2, 2) for the shape parameter; for all other parameters, we used the brms defaults.

We then use the fitted models to estimate using g-computation:^25^

1. The average treatment effect (ATE) in the outcome had each included individual taken a different dose compared to the average total dose (52 mg/kg).
2. The counterfactual outcomes that would have occurred had individuals taken a different dose and been infected with a strain with a different MIC.

We also examined post-treatment stool positivity. Additional methods and results for this outcome are described in Text S1.

### Combining model predictions with %API data

To understand the effect of the observed distributions of %API, we combined our counterfactual predictions with the distributions of %API for ofloxacin and ciprofloxacin found in the medicine quality surveys. To do this, we first set all parameters other than dose and MIC to their average values. Then we used our model to estimate the change in the average outcome when individuals received medicines with a range of %APIs present compared to if they received medicines with 100% API:

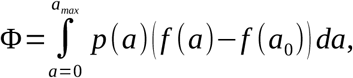

where *a* represents dose, *a_0_* represents a dose with 100% API, *p*(*a*) represents the probability density of each dose (derived from the %API distributions from the literature and the dose that was set at 100%) and *f*(*a*) represents the outcome of interest (fever clearance time or treatment failure probability). We estimate Φ for a range of MICs and values of *a_0_*,

### Alternative models

To examine robustness of modelling choices, we fitted three alternative models consistent with our causal assumptions (detailed in Text S2 and Tables S2).

### Data and code availability

Code and synthetic data to run all analyses are available at https://github.com/scavany/typhoid_sf_antibiotics. Trial data is accessible upon suitable request.

### Ethics

This study represents a secondary analysis of anonymised data from seven trials and publicly available data on medicine quality. Patients or a parent or guardian, for children, gave informed verbal consent before entry into the original trials. All the randomised controlled trials received ethical approval from the Scientific and Ethical Committee of the participating hospital; details are given in the original publications.

## Results

### 1. Distributions of %API

We found one study that surveyed both ciprofloxacin and ofloxacin, and a further six studies surveying only ciprofloxacin (Fig 3), published between 2012 and 2022. These seven studies were from eight different countries, spanning three continents (Mexico, Ghana, Nigeria, Cameroon, Democratic Republic of Congo, Ethiopia, Bangladesh, and Lao PDR). These studies reported %API of 106 samples of ciprofloxacin and 68 of ofloxacin. The median %API for ciprofloxacin was 96.1% (Q1–Q3: 92.1–100%), with 16 samples (15%) falling outside International Pharmacopoeia^26^ tolerance limits of 90–110% (Fig 3). Three samples from one study in Bangladesh^27^ contained less than 60% API. For ofloxacin, the median %API was 103% (Q1–Q3: 98.1–106%), with 91% of samples within 90–110% and none <90%. All ofloxacin samples came from one study in Lao PDR.^28^

**Fig 3:**
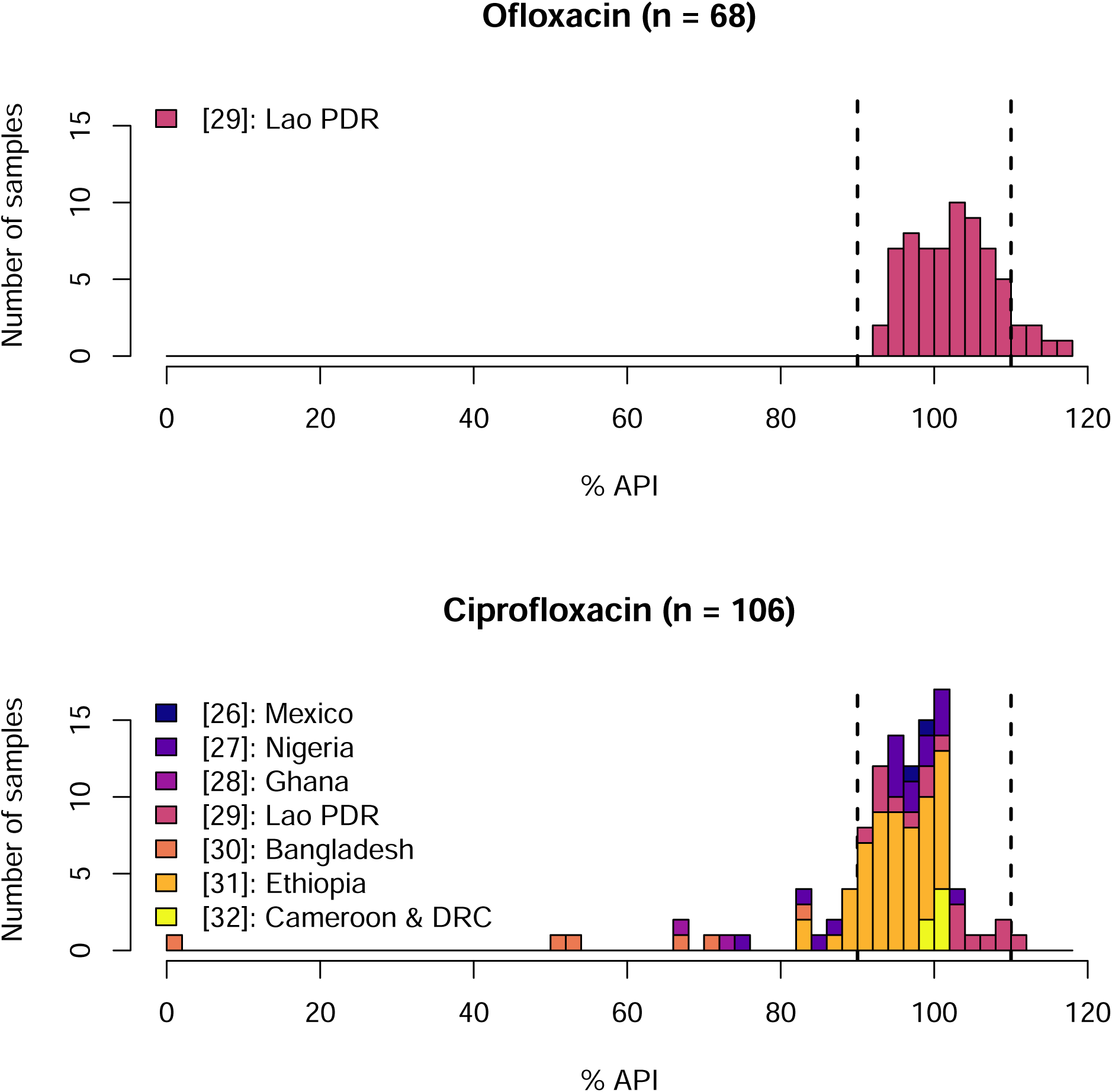
Distribution of the %API for ciprofloxacin (top panel) and ofloxacin (bottom panel) collected in random surveys.^27–33^ The dashed lines represent the tolerance limits for ciprofloxacin tablets as stated by the International Pharmacopeia (12^th^ Edition, 2025).^26^

### 2. Individual-level effects of the incorrect level of API

After excluding 34 patients without MIC data and two without dosing data, 536 patients were included in the analysis. Of these, 104 patients (19.4%) had infections with non-susceptible strains (MIC>0.125 mg/L). The mean fever clearance time was 114 h with a standard deviation of 70.2 h and 72 patients (13.4%) experienced treatment failure. The mean total dose allocated was 51.4 mg/kg (range: 13.8–172 mg/kg).

Using generalized additive models and g-computation, we estimated average treatment effects of different doses compared to the average dose (52 mg/kg). The analysis showed no clear relationship between ofloxacin dose and fever clearance time or treatment failure probability within the dose range of the trials (Fig 4). When exploring how different levels of resistance modified dose effects, both outcomes showed no relationship with ofloxacin dose at MICs deemed susceptible to ofloxacin (i.e., MIC ≤ 0.125 mg/L) by the Clinical and Laboratory Standards Institute (CLSI) (Figs 5 and 6). At higher MICs (e.g., 1 mg/L), however, we observed improved outcomes as dose increased, particularly for fever clearance time (Fig 5). For patients whose infecting strain had an MIC of 1 mg/L, doubling the baseline dose from 52 mg/kg to 104 mg/kg reduced expected fever clearance time by 103h [95% CrI: 21.7–210]. The effect was weaker and more uncertain for failure probability (Fig 6).

**Fig 4.**
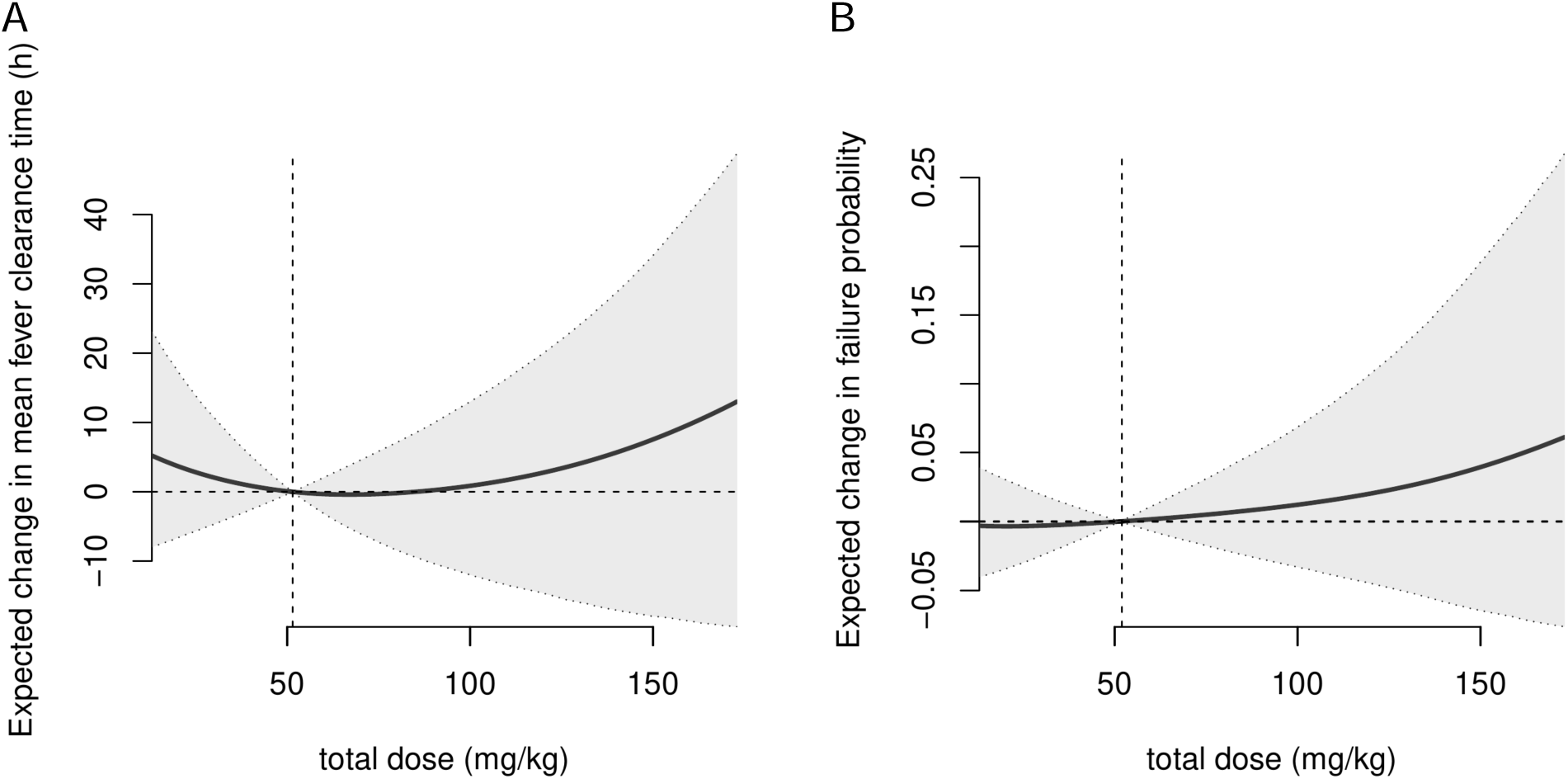
Average treatment effect of different ofloxacin doses on fever clearance time (A) and failure probability (B) compared to a baseline total dose of 52 mg/kg. Effects were only estimated within the range of doses present in the data (13.8–172.4 mg/kg). All other parameters were kept at their baseline values for each individual. The grey shaded region represents a 90% credible interval. The horizontal dashed line represents no change from baseline. The vertical dashed line represents the average dose.

**Fig 5.**
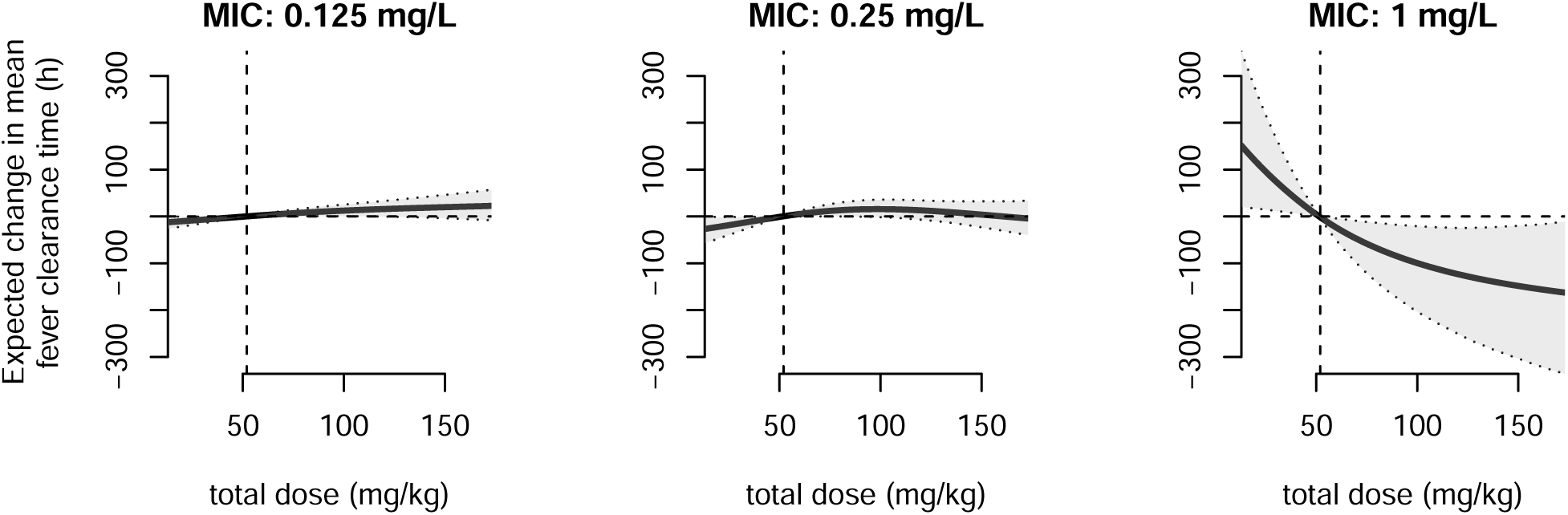
Average treatment effect of different doses of ofloxacin at different MICs (panels) on fever clearance time compared to the average total dose of 52 mg/kg (vertical dashed line). Effects are only shown within the range of doses present in the data (13.8–172.4 mg/kg). All other parameters were kept at their baseline values for each individual. The grey shaded region represents a 90% credible interval. The horizontal dashed line represents no change compared to the average dose.

**Fig 6.**
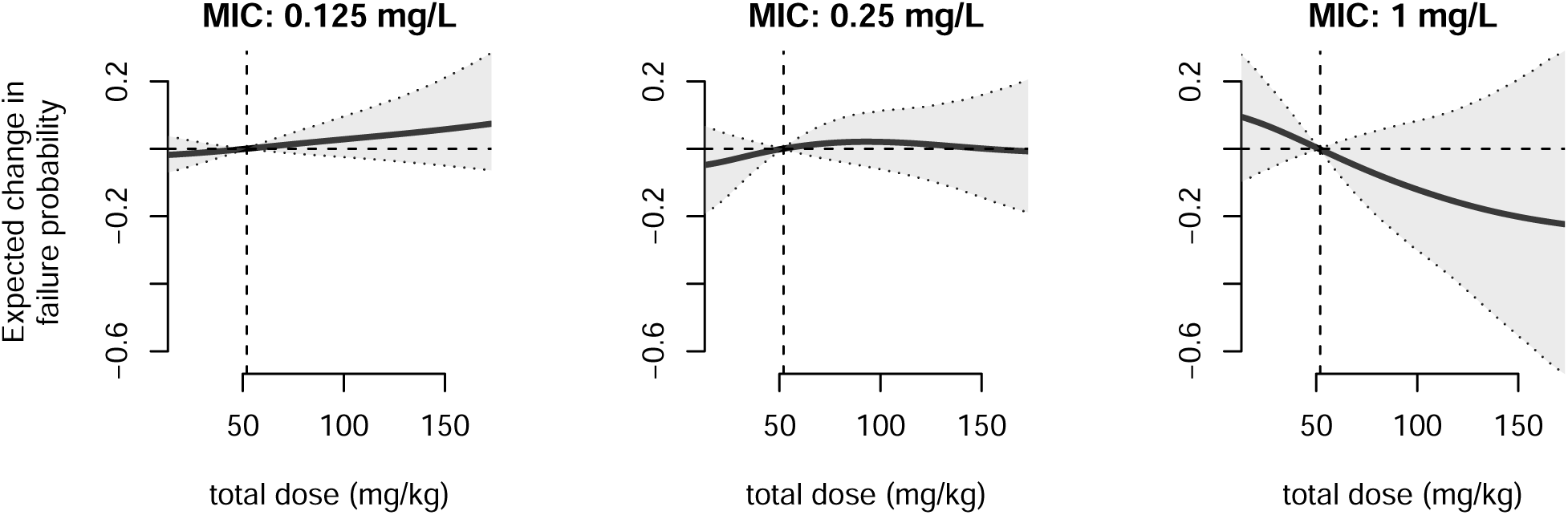
Average treatment effect of different doses of ofloxacin at different MICs (panels) on failure probability compared to the average total dose of 52 mg/kg (vertical dashed line). Effects are only shown within the range of doses present in the data (13.8–172.4 mg/kg). All other parameters were kept at their baseline values for each individual. The grey shaded region represents a 90% credible interval. The horizontal dashed line represents no change compared to the average dose.

### 3. Population-level effects of the incorrect level of API

When ofloxacin contained a distribution of different %APIs according to those in Fig 3, predicted population-level effects differed qualitatively depending on which distribution (Figs 7 and 8). When %API followed the ofloxacin distribution (mean>100%; Fig 3A),^28^ we estimated a 4.1h (90% CrI: 0.75–8.7) reduction in fever clearance time for non-susceptible infections with an MIC of 1 mg/L compared to ofloxacin with 100% API (Fig 7). When %API followed the ciprofloxacin distribution (mean<100%; Fig 3B), we estimated an 11h (90% CrI: 1.9–25) increase in fever clearance time for an MIC of 1 mg/L. When we instead assumed that %API followed the Bangladesh ciprofloxacin distribution (lowest mean %API; Fig 3B),^27^ we estimated a 52.6h (95% CrI: 8.84–116) increase in fever clearance time (Fig S2). These findings are largely robust to the target dose administered (compare panels in Fig 7), but there is greater uncertainty at higher target doses because in that case medicines with the incorrect %API require greater extrapolation from the range of the data.

**Fig 7.**
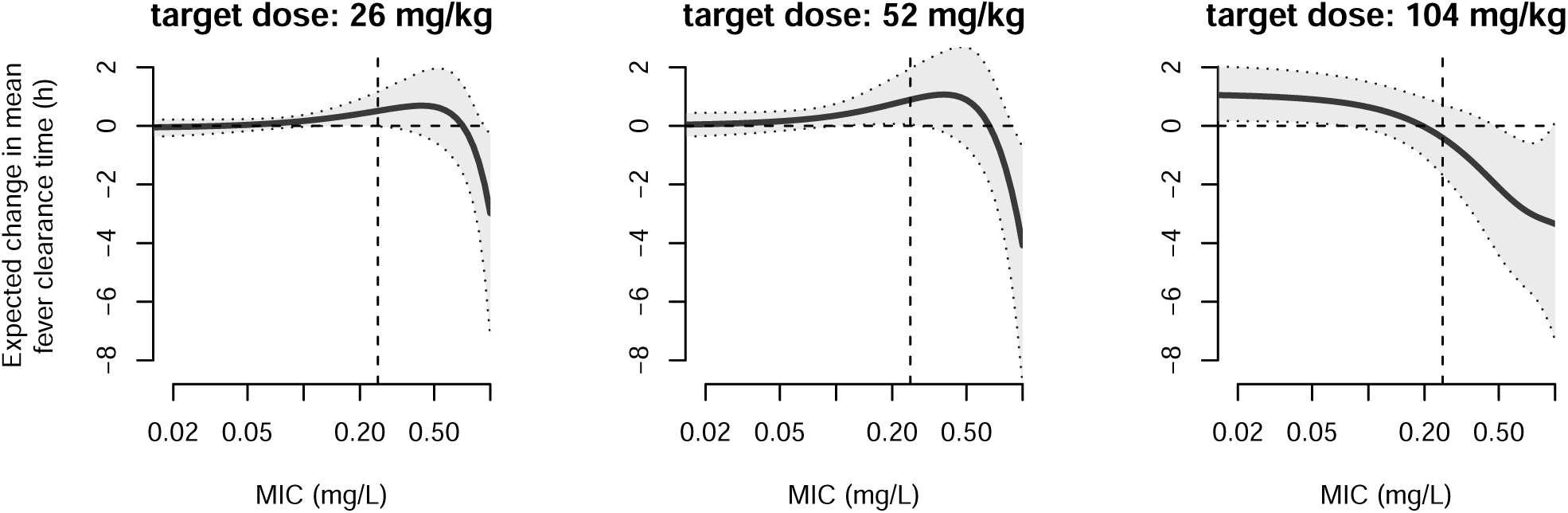
Expected change in fever clearance time when patients take ofloxacin with %API distributed according to the ofloxacin distribution shown in Fig 3A compared to when they take ofloxacin with 100% API. Negative values indicate that the distribution decreased (i.e., improved) the fever clearance time. The shaded region shows 90% credible intervals. The panels show different target doses (i.e., the dose represented by 100% API). The vertical dashed line indicates the breakpoint for ofloxacin; MICs to the right of this line are considered non-susceptible.

**Fig 8.**
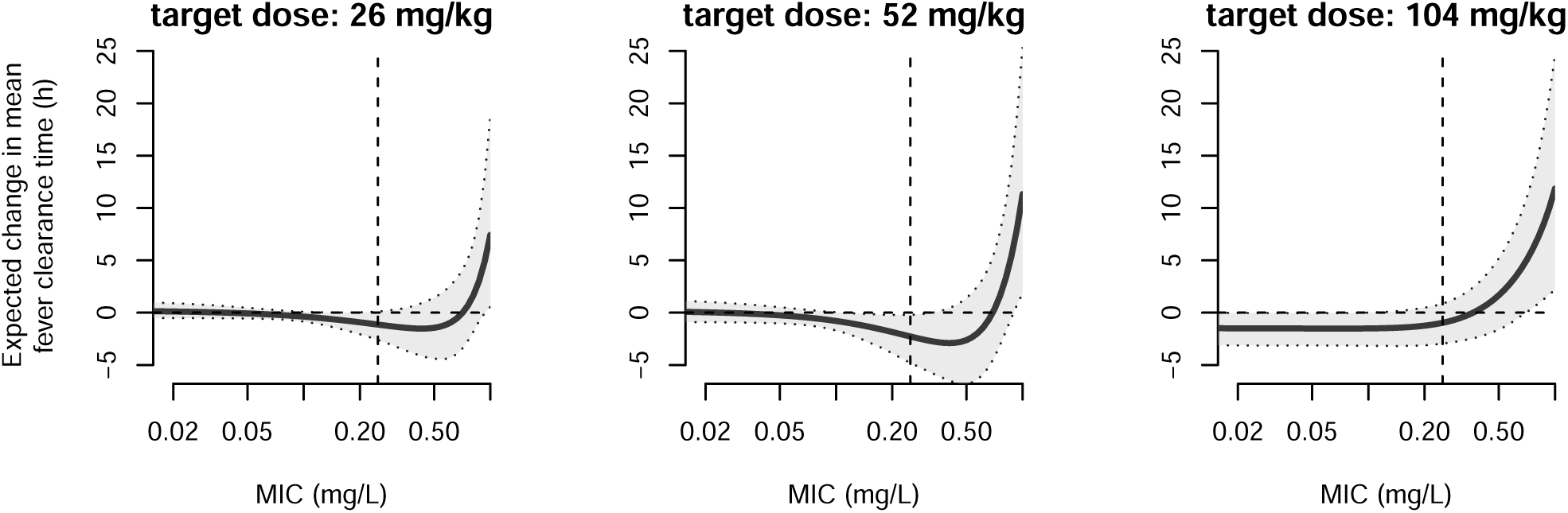
Expected change in fever clearance time when patients take ofloxacin with %API distributed according to the ciprofloxacin distribution shown in Fig 3B compared to when they take ofloxacin with 100% API. Negative values indicate that the distribution decreased (i.e., improved) the fever clearance time. The shaded region shows 90% credible intervals. The panels show different target doses (i.e., the dose represented by 100% API). The vertical dashed line indicates the breakpoint for ofloxacin; MICs to the right of this line are considered non-susceptible.

Population level changes in failure probability were small—when the prescribed dose was 52 mg/kg and the MIC 1 mg/L, we estimated a 0.40 percentage point (pp; 90% CrI: −0.31–1.1) reduction for the ofloxacin distribution (Fig S3), a 0.89pp (90% CrI: −0.70–2.5) increase in the failure probability for ciprofloxacin distribution (Fig S4), and a 4.31pp (95% CrI: −3.38–12.1) increase for the Bangladesh ciprofloxacin distribution (Fig S5).

### Alternative models

Results were largely robust to alternative modelling choices, with Models S1 and S2 showing similar dose–MIC interactions. Our baseline model had the highest predictive skill as measured by the leave-one-out information criterion (Table S2). See Text S2 and for further details on the alternative models.

## Discussion

In this study, we developed a novel framework for estimating clinical impacts of antimicrobials with incorrect %API and applied it to ofloxacin treatment for typhoid. We found that insufficient doses substantially worsen individual-level outcomes when infecting strains have high MICs (e.g., 1 mg/L), particularly for fever clearance time. In the included random surveys of ciprofloxacin and ofloxacin medicine quality, the %API distributions for ciprofloxacin and ofloxacin are both clustered around 100%. Combining these distributions with our individual-level causal effects, the largest detrimental population-level impact we predicted was a mean 11h (90% CrI: 1.9–25) increase in fever clearance time, which occurred when the %API was distributed according to the ciprofloxacin data (Fig 3B), for individuals prescribed a target dose of 52 mg/kg, and with a non-susceptible infection (MIC of 1 mg/L). However, some individual studies found substantially lower %APIs,^27,31^ and in these contexts population-level impacts would be much worse.

Our results indicate that while the effect of SF antibiotics may be small when most strains are susceptible, outcomes can be made substantially worse by the SF antibiotic in settings where non-susceptible or resistant strains dominate. For example, for every patient receiving ofloxacin with 50% API and infected with *S.* Typhi with an MIC of 1 mg/L, fever clearance would take an additional 94h (95% CrI 15–210) compared with if they had received 100% API ofloxacin. Such fever prolongation has great clinical and economic significance, both for the patient and the health care facilities. Our results also imply that it is in locations where MIC is known to be particularly high, such as South Asia,^34^ that we should be particularly vigilant in monitoring the quality of typhoid medications, as these locations may be most at risk of worse outcomes caused by SF antibiotics. The potential risks of SF fluoroquinolones are also likely to have increased over time as the fluoroquinolone MIC of *S.* Typhi increases.^4^ Notably, although *S.* Typhi strains with ofloxacin MIC > 1 mg/L were not observed in the included trials, such high MICs are now common in South Asia.^4,35^

Our finding that the ofloxacin dose impacts outcomes only at higher MICs aligns with the original trials,^12,14,15,17,18^ which found no effect of dose or duration on fever clearance time or treatment failure. It also aligns with Parry’s observation of an inverse relationship between dose/MIC and fever clearance time.^36^ We found a similar inverse relationship between dose/MIC and fever clearance time (Fig S1). Two other studies^4,5^ have also observed that higher MICs were associated with increased fever clearance time.

At least two previous studies have attempted to estimate the public health impact of SF antimicrobials on diseases other than typhoid,^6,9^ both finding that the public health impact of SF antimicrobials could be substantial. A WHO report^6^ estimated that 72,430 childhood pneumonia deaths per year could be attributable to SF antibiotics in their most likely scenario, and both studies^6,9^ independently estimated that around 2–5% of malaria deaths (among under-5s in the case of Renschler *et al.*^9^) could be attributable to SF antimalarials. Unlike these studies, which assumed effects of SF antimicrobials on case fatality risk or drug efficacy, we directly estimated causal effects of incorrect API levels using individual-level patient data.

Our study has at least four limitations that warrant consideration. First, the %API in the antibiotics used in the clinical trials we reanalysed was not reported, and so we implicitly assumed that all contained 100% API. If the distribution instead had a mean lower or higher than 100%, this would shift the dose-response relationships to the left or the right respectively and increase the uncertainty. In either case, assuming medicines were allocated randomly and the distribution of %API did not change substantially between studies, then assuming that all medicines had 100% API is unlikely to have qualitatively changed our findings. Second, we found only one random survey reporting the %API of ofloxacin and thus used ciprofloxacin surveys as a proxy for the ofloxacin distribution. Moreover, only six random surveys reported the %API of ciprofloxacin with fewer than 200 samples in total, spanning three continents and ten years (2012–2022), making it hard to generalize their findings. In places and times where there is a locally high prevalence of very low %API medicines, such as those found in the Bangladesh study,^27^ treatment outcomes may be substantially worse. Third, we focused solely on the impact of the API concentration, ignoring other mechanisms by which SF medicines could affect health, such as the toxic effects of medicines containing harmful ingredients^37^ or too much API, reduced bioavailability,^38^ or the potential for SF antibiotics to affect antimicrobial resistance.^38,39^ Finally, extrapolation beyond the 13.8–172 mg/kg dose range may be inappropriate as such doses were not received by any patients in the trials. This is pertinent when considering the %API distribution of ciprofloxacin, for which a small number of medicines contained very little API. Notably, estimates of average fever clearance time from the pre-treatment era of 756h are substantially longer than would be obtained from extrapolating any of our models to a dose of 0, suggesting that medicines containing 0% API may have worse outcomes than implied by our results.^40^

Our framework for quantifying the impact of SF antimicrobials demonstrates that the population-level impacts of SF antimicrobials on typhoid outcomes may be heightened when the MIC of *S.* Typhi is high and the SF antimicrobials have low %API. This is a situation that has become more common as fluoroquinolone MICs for *S.* Typhi have risen.^4,35^ This framework could be applied to other antimicrobial–pathogen pairs for which there are data to inform the distribution of %API and the dose-response relationship for key clinical outcomes. Future work should include improving our understanding of the %API distribution in different settings and how SF antimicrobials affect bioavailability, for instance through dissolution testing. It is also critical that future studies address the role of SF antimicrobials in driving antimicrobial resistance.

## Acknowledgements

We thank the wider FORESFA Collaboration for their advice and support for this work. We also thank all of the individuals involved in conducting the original studies whose data we utilized and all of the patients and their families who participated in the trials.

## Transparency Declaration

None to declare.

