## Supplementary material for "A novel framework for quantifying the clinical impact of substandard and falsified antimicrobials: application to typhoid fever": Text S1

*Text S1: additional methods and results for stool positivity outcome*

*Additional methods.* Stool samples that were positive following the completion of treatment could give an indication of the potential for onward transmission. For this outcome, 88/572 (15%) patients did not have a stool sample taken and so were removed from the analysis. For a further 67 (12%) patients the number of positive stool samples taken was not recorded, and instead it was recorded whether at least one sample was positive. For all included patients the total number of stool samples tested was recorded. With this in mind, we used a mixed likelihood function, that is,

$$\mathscr{L}(p_i|n_i, r_i) = \binom{n_i}{r_i} p_i^{r_i} (1-p_i)^{n_i - r_i}$$

if $r_i$ is known, and

$$\mathscr{L}(p_i|n_i, r_i > 0) = 1 - (1-p_i)^{n_i}$$

if $r_i$ is unknown, where $r_i$ represents the number of positive stool samples, $n_i$ the total number of stool samples, and $p_i$ the probability of a positive stool sample predicted by the model and its parameters.

*Results.* To investigate the effect of antibiotic dose on potential for onward transmission, we also examined an additional outcome: stool positivity following the completion of treatment. Excluded patients had a higher average total dose than included patients (54.2 mg/kg (90% CI: 51.2 – 57.3) *vs.* 39.6 mg/kg (90% CI: 34.6 – 44.6) respectively) and a higher average MIC (0.178 mg/L (90% CI: 0.156 – 0.200) *vs.* 0.0879 mg/L (90% CI: 0.0620 – 0.114)). Due to the reduced data set and relatively low number of positive samples, it was difficult to conclude anything about the effect of dose on stool positivity (Fig S1), although the trend was in the direction of improved outcomes at higher doses when the infecting strain had a high MIC.

*Text S2: Additional details on alternative models*

*Methods.* To examine the robustness of our results to our modelling choices, we also undertook the analysis with three alternative models, each of which are consistent with the causal assumptions described in Fig 2. These alternative models are listed in Table S2. Briefly, in Model S1 we divided daily dose by MIC prior to inclusion in the model, and included an interaction between this term and duration of treatment; in Model S2 we included a three-way interaction between daily dose, MIC, and duration of treatment; in Model S3 we included interactions between dose and duration of treatment and dose and weight, but excluded MIC from the model.

*Results and discussion.* For Model S1 we divided daily dose (mg/kg/day) by MIC (mg/L) before including it in the model and include an interaction between this term and duration of treatment. In this case, increasing dose/MIC led to clear reductions in both fever clearance time and treatment failure probability at all treatment durations (Figs S6 and S7). The effect was particularly strong when dose/MIC was high (i.e., when dose is low and MIC high), which is commensurate with our baseline finding that lower doses led to worse outcomes when MIC was high. Model S2, which includes a three-way interaction between daily dose, MIC, and treatment duration, showed a decrease in failure probability at high doses, but did not show a relationship between fever clearance time and dose at any MIC (Figs S8 and S9), counter to our findings with the baseline model and model S1. However, model S2 had a worse predictive performance than the baseline model on fever clearance time, as approximated by the leave-one-out information criterion (LOOIC; Table S3). Model S3, which does not include MIC, has no clear relationship between dose and the outcomes of interest (S10 and Fig S11s). In all three of these alternative models there is little evidence of an interaction between duration and dose. Comparing all models' predictive performance using LOOIC, the baseline model and model S1 had the best predictive performance on fever clearance time, the baseline model and model S2 had the best predictive performance on treatment failure, and model S3 was worst on both outcomes (Table S3)

*Table S1: Summary of included trial arms. Details are given only for ofloxacin arms. Where more than one arm of a study included ofloxacin treatment, the study has two rows; the arms can be distinguished by differing durations of treatment allocated.*

| **Journal article** | **Treatment Duration (days)** | **Ofloxacin Dose** | **Number of patients** | **Sex** (% female) | **Age** (years) median (IQR) | **Bodyweight** (kg) median (IQR) | **MIC** (mg/L) median (IQR) |
|---|---|---|---|---|---|---|---|
| Smith *et al.*, *Antimicrobial Agents and Chemotherapy* (1994)[16] | 5 | 400 mg/day | 19 | 31.6 | 21 (18–23) | 46.0 (36.0–48.5) | 1/32 (1/32–1/32) |
| Vinh *et al.*, *Antimicrobial Agents* | 3 | 15 mg/kg/day | 52 | 48.1 | 9 (5–12) | 18.4 (13.0–24.6) | 1/16 (1/32–1/16) |
| *and Chemotherapy* (1996)[13] | 2 | 15 mg/kg/day | 50 | 40.0 | 8 (6–11) | 19.8 (14.5–23.9) | 1/16 (1/32–1/16) |
| Chinh *et al*. *Transactions of the* | 3 | 10 mg/kg/day | 55 | 52.7 | 24 (18.5–27.5) | 45.0 (41.0–48.0) | 1/16 (1/32–1/16) |
| *RSTMH* (1997)[17] | 2 | 15 mg/kg/day | 46 | 65.2 | 23 (19–30) | 46.0 (43.0–48.0) | 1/16 (1/32–1/16) |
| Cao *et al*. *The Pediatric Infectious Disease Journal* (1999)[14] | 5 | 10 mg/kg/day | 68 | 44.1 | 6.5 (5–10) | 17.0 (14.0–20.3) | 1/16 (1/16–1/16) |
| Chinh *et al*. *Antimicrobial Agents and Chemotherapy* (2000)[18] | 5 | 400 mg/day | 45 | 44.4 | 24 (19–28) | 47.0 (44.0–54.0) | 1/8 (1/32–1/2) |
| Vinh *et al*. *Annals of Tropical* | 3 | 10 mg/kg/day | 89 | 44.9 | 8 (6–10) | 17.0 (13.5–20.0) | 1/16 (1/32–1/16) |

| | | | | | | | |
|---|---|---|---|---|---|---|---|
| *Paediatrics* (2005)[12] | 2 | 10 mg/kg/day | 77 | 42.9 | 8 (6–11) | 17.0 (14.0–21.0) | 1/16 (1/16–1/16) |
| Parry *et al*. *Antimicrobial Agents and Chemotherapy* (2007)[15] | 7 | 20 mg/kg/day | 69 | 49.2 | 8 (7–11) | 20.0 (15.0–24.0) | 1 (1/2–1) |

*Table S2. Alternative models.*

| Model | Equation | Description |
|---|---|---|
| S1 | `outcome ~ t2(dose_over_MIC, duration) + s(wt) + s(study, bs = “re”)` | Divide dose by MIC. Includes interaction between this value and treatment duration. |
| S2 | `outcome ~ t2(dose, duration, MIC) + s(wt) + s(study, bs = “re”)` | Three-way interaction between dose, treatment duration, and MIC. |
| S3 | `outcome ~ t2(dose, duration, wt) + s(study, bs = “re”)` | Three-way interaction between dose, treatment duration, and weight. **No consideration of MIC.** |

*Table S3. Models compared by leave-out-out information criterion (LOOIC). By this metric, models with a lower LOOIC are expected to have a greater out-of-sample predictive performance; we display the difference in LOOIC from the best model, and the standard error of the difference. LOOIC is an approximation of leave-one-out cross-validation (LOOCV) using Pareto smoothed importance sampling (PSIS), and is calculated in the brms package.*

| **Model** | **Fever clearance time** | **Failure probability** |
|---|---|---|
| Baseline model | 0.0 | 0.0 |
| S1 model | 6.1 (11.2) | 14.2 (6.4) |
| S2 model | 12.4 (3.7) | 1.6 (4.3) |
| S3 model | 74.8 (19.4) | 22.0 (9.0) |

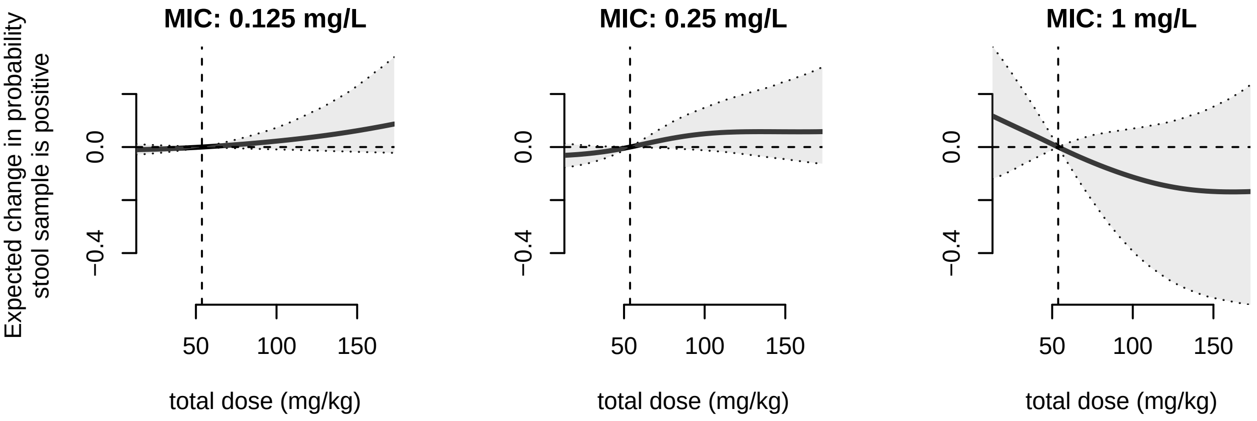


*Fig S1. Average treatment effect of different ofloxacin doses at different MICs (panels) on the probability of a positive stool sample compared to a baseline total dose of 52 mg/kg (vertical dashed line). Effects are only shown within the range of doses present in the data (13.8–172.4 mg/kg). All other parameters were kept at their baseline values for each individual. The grey shaded*

*region represents a 90% credible interval. The horizontal dashed line represents no change from baseline.*

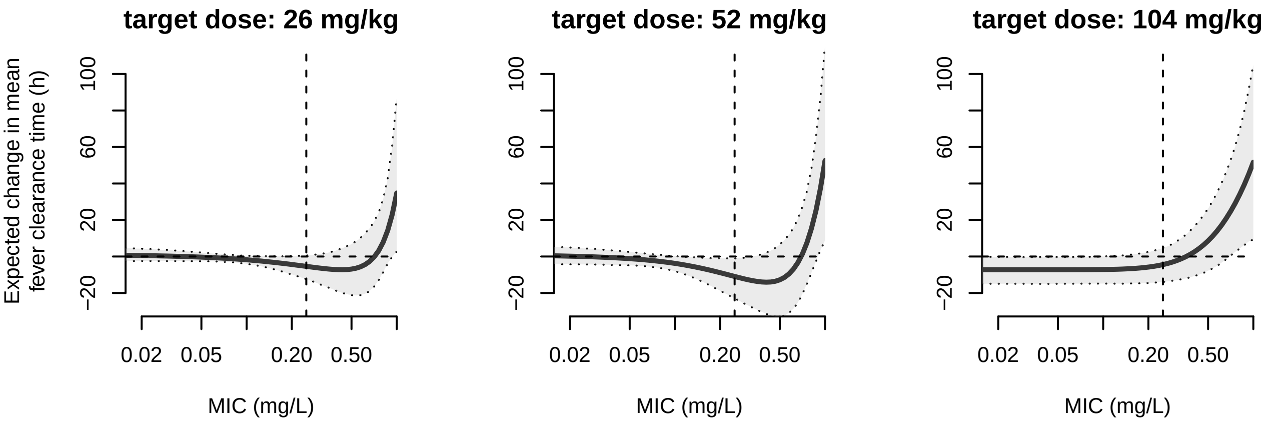


*Fig S2. Expected change in fever clearance time when patients take ofloxacin with %API distributed according to the ciprofloxacin distribution found in Bangladesh[27] shown in Fig 3B compared to when they take ofloxacin with 100% API. Negative values indicate that the distribution decreased (i.e., improved) the fever clearance time. The shaded region shows 90% credible intervals. The panels show different target doses (i.e., the dose represented by 100% API). The vertical dashed line indicates the breakpoint for ofloxacin; MICs to the right of this line are considered non-susceptible.*

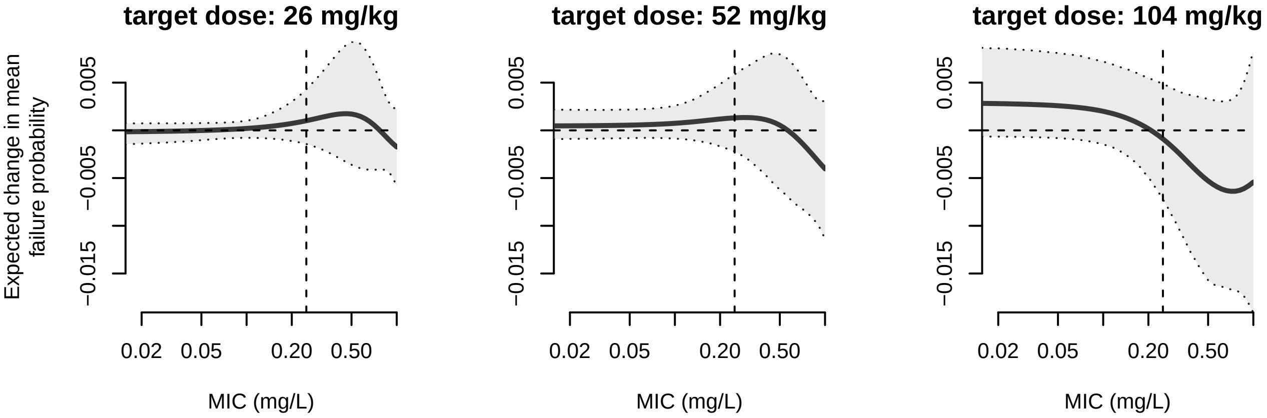


*Fig S3. Expected change in failure probability when patients take ofloxacin with %API distributed according to the ofloxacin distribution shown in Fig 3A compared to when they take ofloxacin with 100% API. Negative values indicate that the distribution decreased (i.e., improved) the fever clearance time. The shaded region shows 90% credible intervals. The panels show different target doses (i.e., the dose represented by 100% API). The vertical dashed line indicates the breakpoint for ofloxacin; MICs to the right of this line are considered non-susceptible.*

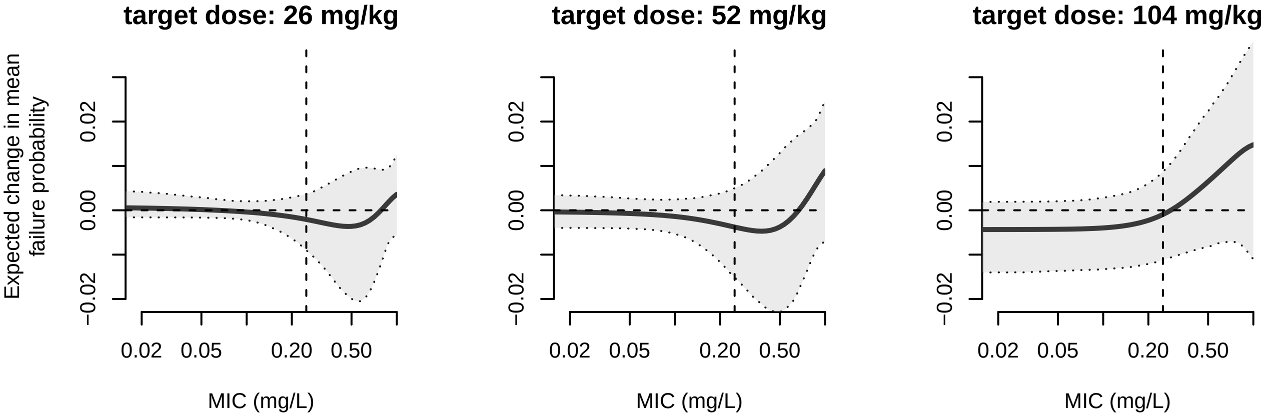


*Fig S4. Expected change in failure probability when patients take ofloxacin with %API distributed according to the ciprofloxacin distribution shown in Fig 3B compared to when they take ofloxacin with 100% API. Negative values indicate that the distribution decreased (i.e., improved) the fever clearance time. The shaded region shows 90% credible intervals. The panels show different target*

*doses (i.e., the dose represented by 100% API). The vertical dashed line indicates the breakpoint for ofloxacin; MICs to the right of this line are considered non-susceptible.*

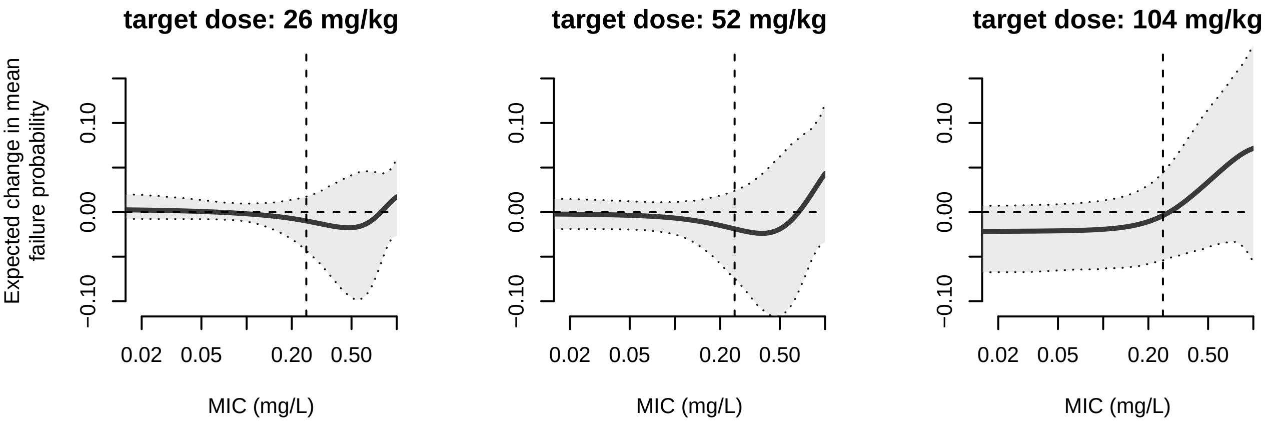


*Fig S5. Expected change in failure probability when patients take ofloxacin with %API distributed according to the ciprofloxacin distribution found in Bangladesh[27] shown in Fig 3B compared to when they take ofloxacin with 100% API. Negative values indicate that the distribution decreased (i.e., improved) the failure probability. The shaded region shows 90% credible intervals. The panels show different target doses (i.e., the dose represented by 100% API). The vertical dashed line indicates the breakpoint for ofloxacin; MICs to the right of this line are considered non-susceptible.*

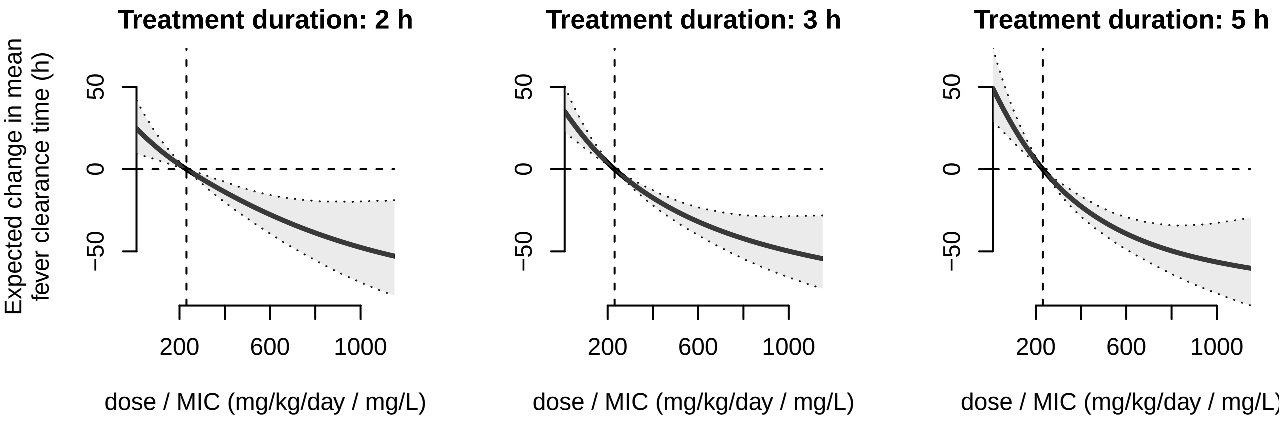

*Fig S6. Average treatment effect of different ofloxacin dose / MICs at different treatment durations (i.e., patients received the dose shown on the x-axis for the duration shown above the panels) on fever clearance time compared to a baseline dose / MIC of 230 mg/kg/day / mg/L (vertical dashed line) using model S1. Effects are only shown within the range of doses present in the data (13.8–172.4 mg/kg). All other parameters were kept at their baseline values for each individual. The grey shaded region represents a 90% credible interval. The horizontal dashed line represents no change from baseline.*

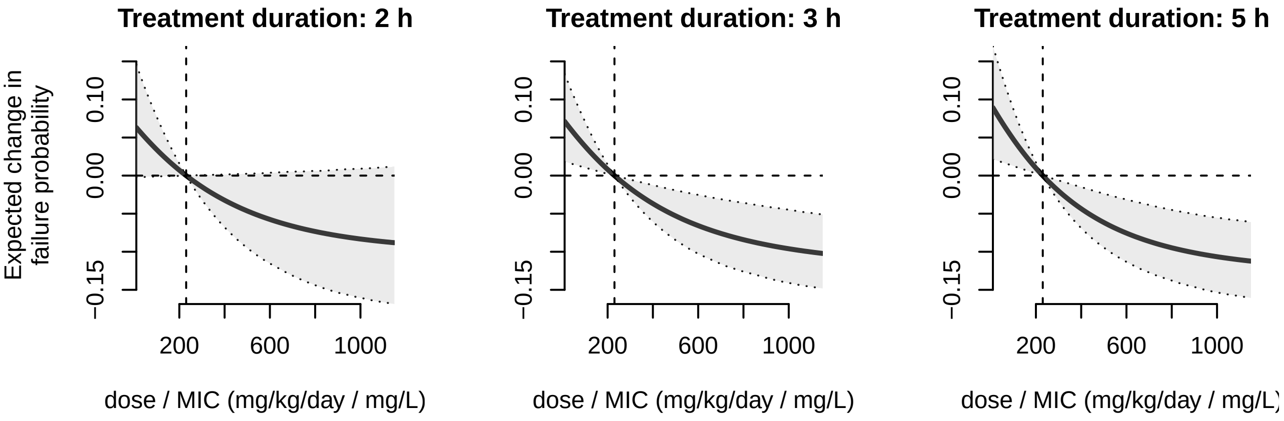


*Fig S7. Average treatment effect of different ofloxacin dose / MICs at different treatment durations (panels) on failure probability compared to a baseline dose / MIC of 230 mg/kg/day / mg/L (vertical dashed line) using model S1. Effects are only shown within the range of doses present in the data (13.8–172.4 mg/kg). All other parameters were kept at their baseline values for each individual. The grey shaded region represents a 90% credible interval. The horizontal dashed line represents no change from baseline.*

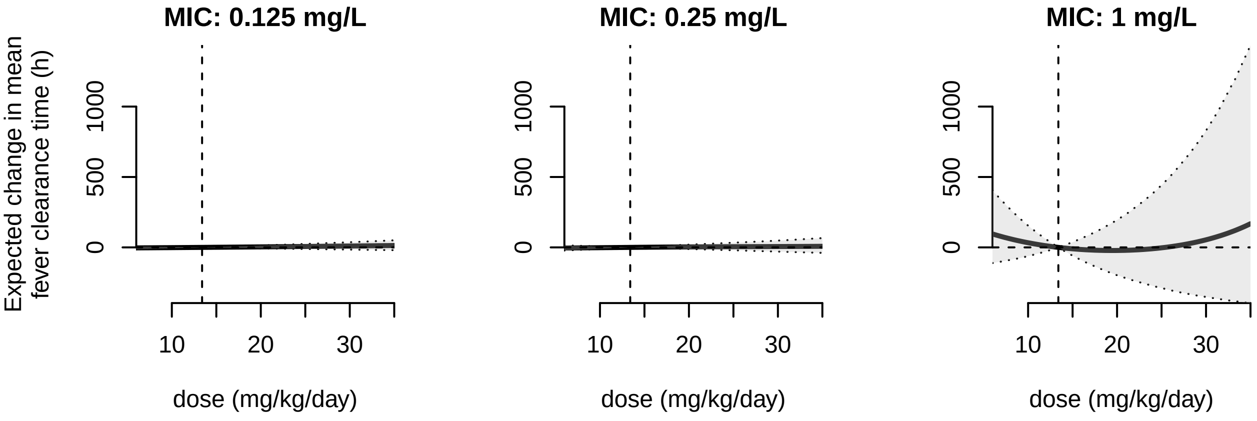


*Fig S8. Average treatment effect of different ofloxacin doses at different MICs (panels) on fever clearance time compared to a baseline dose of 14 mg/kg/day (vertical dashed line) using model S2. Effects are only shown within the range of doses present in the data (13.8–172.4 mg/kg). All other parameters were kept at their baseline values for each individual. The grey shaded region represents a 90% credible interval. The horizontal dashed line represents no change from baseline.*

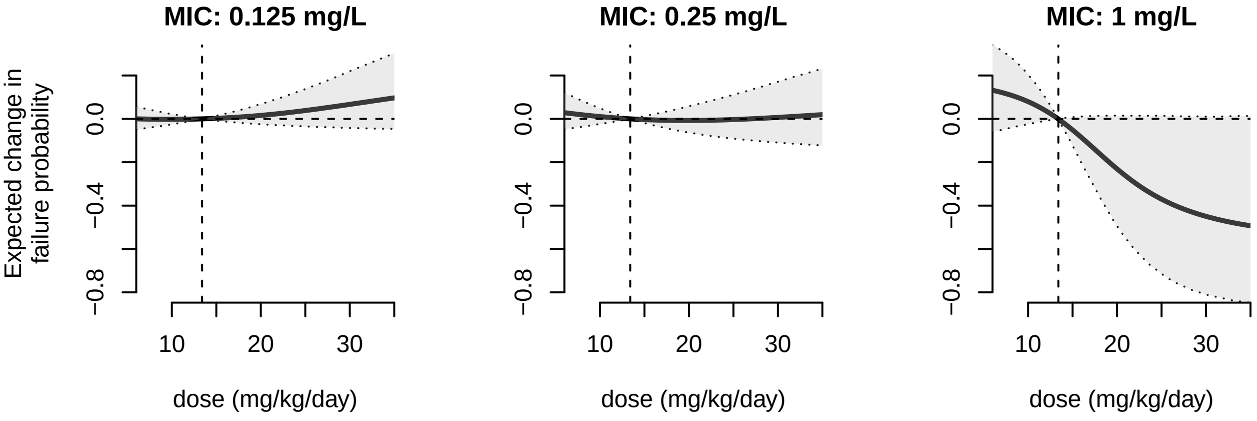


*Fig S9. Average treatment effect of different ofloxacin doses at different MICs (panels) on failure probability compared to a baseline dose of 14 mg/kg/day (vertical dashed line) using model S2. Effects are only shown within the range of doses present in the data (13.8–172.4 mg/kg). All other*

*parameters were kept at their baseline values for each individual. The grey shaded region represents a 90% credible interval. The horizontal dashed line represents no change from baseline.*

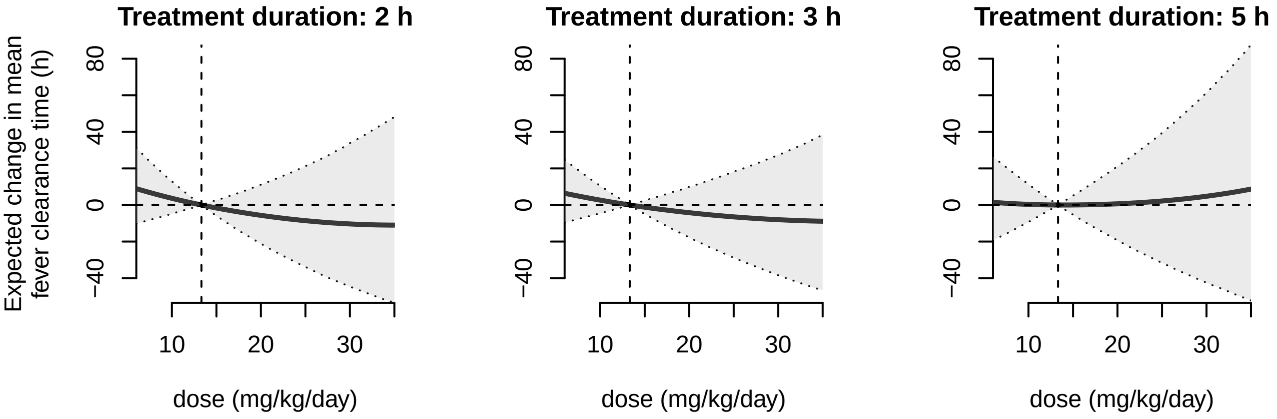


*Fig S10. Average treatment effect of different ofloxacin doses at fever clearance times (panels) on failure probability compared to a baseline dose of 14 mg/kg/day (vertical dashed line) using model S3. Effects are only shown within the range of doses present in the data (13.8–172.4 mg/kg). All other parameters were kept at their baseline values for each individual. The grey shaded region represents a 90% credible interval. The horizontal dashed line represents no change from baseline.*

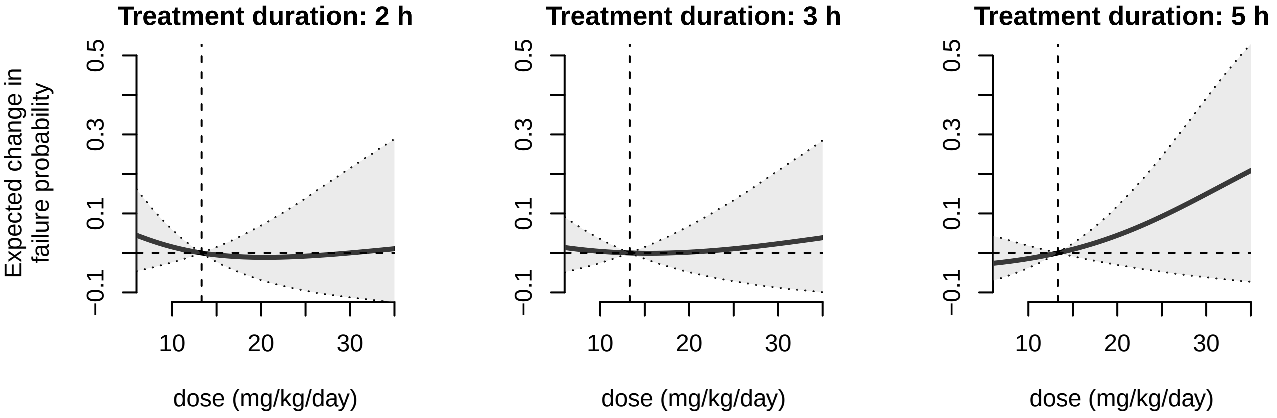

*Fig S11. Average treatment effect of different ofloxacin doses at treatment durations (panels) on failure probability compared to a baseline dose of 14 mg/kg/day (vertical dashed line) using model S3. Effects are only shown within the range of doses present in the data (13.8–172.4 mg/kg). All other parameters were kept at their baseline values for each individual. The grey shaded region represents a 90% credible interval. The horizontal dashed line represents no change from baseline.*
